# Measuring voluntary and policy-induced social distancing behavior during the COVID-19 pandemic

**DOI:** 10.1101/2020.05.01.20087874

**Authors:** Youpei Yan, Amyn A. Malik, Jude Bayham, Eli P. Fenichel, Chandra Couzens, Saad B. Omer

## Abstract

Staying home and avoiding unnecessary contact is an important part of the effort to contain COVID-19 and limit deaths. Every state in the United States enacted policies to encourage distancing, and some mandated staying home. Understanding how these policies interact with individuals’ voluntary responses to the COVID-19 epidemic is critical for estimating the transmission dynamics of the pathogen and assessing the impact of policies. We use the variation in policy responses along with smart device data, which measures the amount of time Americans stayed home, to show that there was substantial voluntary avoidance behavior. We disentangle the extent to which observed shifts in behavior are induced by policy and find evidence of a non-trivial voluntary response to local reported COVID-19 cases and deaths, such that around 45 cases in a home county is associated with the same amount of time at home as a stay-at-home order. People responded to the risk of contracting COVID-19 and to policy orders, though the response to policy orders crowds out or displaces a large share of the voluntary response, suggesting that, during early stages of the U.S. outbreak, better compliance with social distancing recommendations could have been achieved with policy crafted to complement voluntary behavior.

**Significance Statement:** Americans are spending substantially more time at home to reduce the spread of COVID-19. This behavioral shift is a mix of voluntary disease avoidance and policy-induced behavioral changes. Both need to be accounted for. Disentangling voluntary from policy-induced behavioral changes is critical for governments relaxing or renewing restrictions. A substantial share of the behavioral response appears to be voluntary, but this behavior was offset by strong stay-at-home orders. Local testing and rapid reporting is a first step to making better use of voluntary behavioral changes.

## Introduction

Worldwide, people stayed home to reduce transmission of SAR-CoV-2, the virus causing COVID-19 pandemic. Years of theory suggest that people are likely to alter behavior voluntarily to avoid becoming sick, including staying home (1-5). Evidence from the 2009 H1N1 swine flu epidemic (6-8), Lyme Disease (9), the 2003 SARS epidemic (10, 11), HIV (12, 13), and emerging evidence from COVID-19 (14-16) supports the theory that people alter behavior in response to infectious disease risk. Yet, the 2020 COVID-19 epidemic is the first modern epidemic in the United States where most people were ordered by governments to reduce time in public. Therefore, it is one of the first opportunities to investigate how public health orders in the form of non-pharmaceutical interventions interact with voluntary behavioral shifts during epidemics. Recent studies (17-19) have attempted to evaluate the effectiveness of restrictions that force people to avoid leaving home in order to slow and stop the spread of SAR-CoV-2. Each of these studies openly assumes no voluntary change in behavior as the alternative to restrictions. Yet, voluntary behavior is a fundamental part of the transmission process. Fenichel et al. (1) showed that if the true data generating process for an epidemic involves the theoretically predicted voluntary behavioral avoidance response, then an empirical model that assumes away that behavioral response can fit the epidemiological data as well as a model that specifies the true data generating process. The problem is that the misspecified model cannot provide information about behavioral response, whether voluntary or because of public health orders.

It is important to disentangle the roles of voluntary response from policy-induced behavioral change. Moreover, it is important to understand how these two forces interact. This information is critical for assessing the epidemiological risk and the potential short-run economic benefits of relaxing policies that restrict behavior. It is also important for learning from past policies in the event that public health orders are needed again. If the policy orders reduce or replace voluntary contributions of behavioral change, crowding out, then estimates of the effect of a policy that ignores voluntary behavioral responses should be regarded as an upper bound. Conversely, if public health orders encouraged voluntary responses, then estimates that ignore voluntary behavior underestimate the role of these policies in altering the course of the epidemic.

We focus on the locations where people allocate their time budget, using smart devices as the primary metric of behavior. The smart device data is used as a measure of time allocation. For well over a decade, epidemiologists have used surveys of contact behavior to parameterized epidemiological models (20-24). Bayham et al. (8) refined earlier work by Zagheni et al. (25) to infer likely contact patterns from the American Time Use Survey (ATUS). They showed that time use data, based on the ATUS, could produce similar contact patterns to those based on the detailed surveys epidemiologist relied on. Bayham et al. also showed that in the case of H1N1, conditioning cases on time spent at home gave a similar reduction in cases as an epidemiological model using the fully specified contact structure. Hence, we use the smart device data to measure the time spent at home as a measure of avoidance behavior. We confirm the primary results with other measures of time use.

We define the voluntary response as the marginal response to reported cases after controlling for policy interventions. We focus on reported cases within the county, which provides a relatively local measure of risk. We also consider reports of national and state cases. We focus on three policy orders that led to involuntary behavioral responses. First, we consider stay-at-home. These were often colloquially called “lock-downs.” Second, we consider school closures that induce parents to stay home from work to care for children. Third, we consider the emergency orders that raised awareness and may have led businesses to close or encourage working from home. In the supplemental material, we repeat the analysis using reported deaths instead of, and along with, cases. We acknowledge that the voluntary aspect of behavior is hard to define. Closures likely increased the salience of concern for COVID-19, making “voluntary” difficult to define in the COVID-19 upheaval, which is why we focus on the early phase of the epidemic in the United States. However, the salience and other indirect policy impacts may have been achievable through more targeted and, perhaps, less costly policies.

Crowding out effects are well established in public finance when state spending replaces the private provision of public goods, and this phenomenon also extends to non-monetary contributions. Ostrom (26) summarizes the evidence, writing “external interventions crowd out intrinsic motivation if the individuals affected perceive them [the policies] to be controlling.” She argues that many policies adopted in modern democracies presume authorities must solve all collective action problems, thereby crowding out citizenship, wasting resources and undermining democracy. Lock-down policies seem likely to fit this characterization, and empirical evidence suggests that as mandatory involuntary contributions increase, voluntary contributions are increasingly crowded out, even when there is a private benefit to the contribution (27).

All 50 States and the District of Colombia in the United States issued emergency orders, and 40 (39 States and the District of Colombia) issued stay-at-home or shelter-in-place orders, which are essentially two names for the same thing (hereafter stay-at-home). However, there is substantial heterogeneity in the timing of local cases and stay-at-home orders (Figure 1), but most counties were under emergency order prior to experiencing a case (Figure S1).

**Figure 1.**
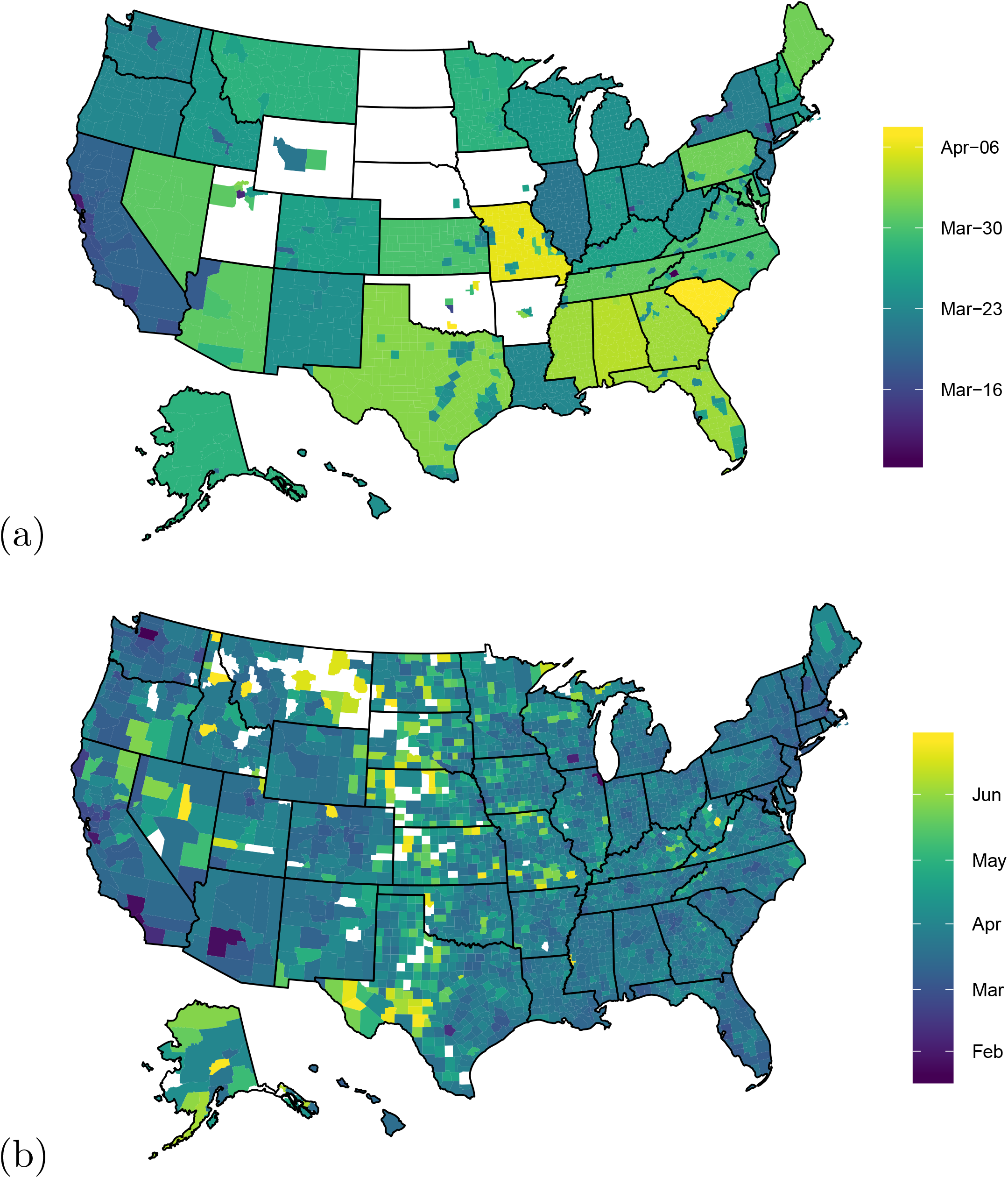
(a) Date that counties enacted stay-at-home order and (b) date of the first case reported within a county.

Here, we use the variation in policy responses along with smart device data to measure the amount of time Americans stayed home, and adjusted other behaviors, in response to pathogen risk and stay-at-home orders. We contribute to the body of evidence that finds strong voluntary avoidance behavior. We also contribute to the body of evidence that finds a strong effect of policy. We fill a gap in these literatures by connecting them and find evidence of a negative interaction, implying that policies that restrict behavior crowd out the voluntary response to pathogen risk.

## Results

We use SafeGraph smart device median home dwell time data at the Census block group level averaged to the county level for all counties in the United States and Washington DC between January 31, 2020, and April 21, 2020, which covers two weeks before the earliest declaration of emergency and two weeks after the latest stay-home policy during the first phase of the US epidemic. Of the 3,104 US counties, 1,944 are metropolitan counties, which generally have a greater share of urban and suburban communities than the non-metropolitan counties. The daily mean of the median time spent at home on January 31 was 576.8 minutes (sd 86.8). Time at home gradually declined from January 31, reaching a minimum of 411.8 minutes (sd 99.5) on February 25. Time spent at home then increased, peeking on April 12, 923.5 minutes (sd: 158.2) (Figure 2). The SafeGraph retail visitation data suggest that people increased visits to stores like Costco, Walmart, and Target around the weekend that preceded a major wave of state stay-at-home orders, perhaps in anticipation of restrictive policies (28).

**Figure 2.**
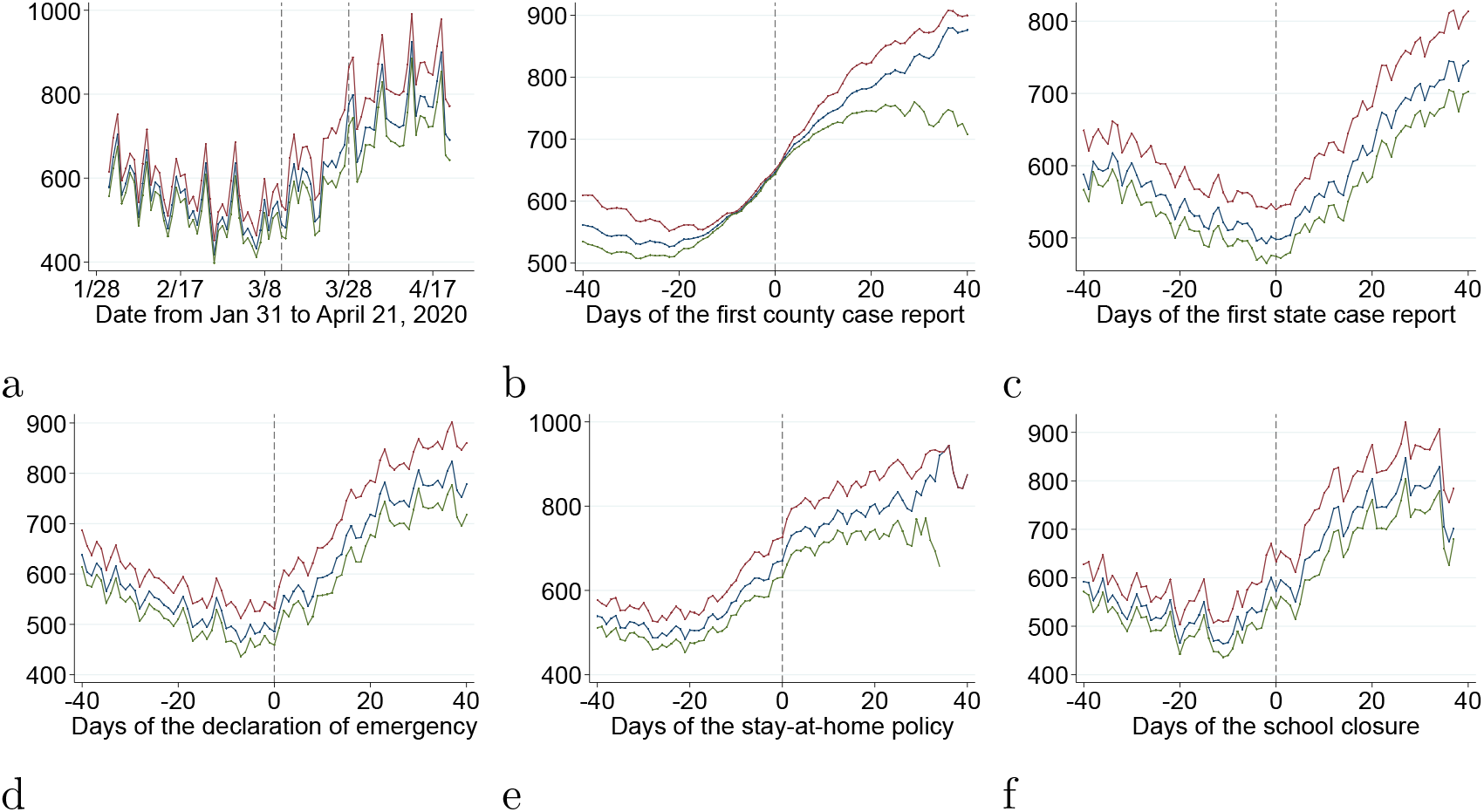
Shows trends for the mean time at home in minutes for the full sample (blue line), metropolitan counties (red line), and non-metropolitan counties (green line) by (a) calendar date with vertical lines showing the median date for emergency orders and stay-at-home policies, (b) aligned to the first case reported at the county-level, (c) aligned to the first case reported at the state-level, (d) aligned to the emergency declarations, (e) aligned to school closure, and (f) aligned to the stay-at-home policy.

Time spent at home was at its lowest level and began rising prior to emergency declarations (Figure 2). This adds to the evidence that a portion of the behavioral response was voluntary. Median time at home continued to increase following the emergency declaration.

We use a dataset of 251,992 observations to estimate a regression model of the log of time spent at home on the log of reported local and national cases, the presence of distancing policies, and the interaction between reported county cases and stay-at-home orders to disentangle voluntary avoidance behavior from the effect of policies and investigate if policy orders crowd out voluntary responses (see Methods for specification details). Almost all coefficients were precisely estimated, and regression tables for all specifications are provided in the SI.

Americans increased time at home in direct response to case reports, death reports, school closures, and stay-at-home orders. The voluntary response appears to be partially crowded out and offset by policies (Table 1). We find that a one percent change in cumulative reported cases in a county yields approximately a 4 percent increase in time spent at home, which is stable across specifications but is reduced slightly when state and county cases are included in the regression. However, we also find that a one percent increase in state-level cases is associated with approximately a 4 percent increase in time at home. Instead, focusing on new cases, we find that a one percent increase in new county (state) cases yields approximately a 2 (3) percent increase in time at home. We find that a one percent increase in reported county or state deaths yields a 4-6 percent increase in time at home. School closures increase time at home by approximately 8 to 16 percent. Stay-at-home orders increased time at home by 8 to 23 percent. The interaction between stay-at-home orders and cumulative cases and deaths is robustly negative and always precisely estimated, except when interactions between county and state cases are included in the same regression (Table 2). In those cases, both coefficients are negative, and at least one is precisely estimated. This is evidence in support of the hypothesis that stay-at-home policies crowded out some of the voluntary response. For the primary specification, the interaction term is greater than half the size of the voluntary response, thereby crowding out a substantial share of the voluntary effort, and this goes up when including day-fixed effects.

**Table 1:** Time at home associated with county-case reports under the base model, with epidemic-day (based on the first county-case report) fixed effects, day since stay-at-home order fixed effects, and with border states’ case report (Jan 31 – April 21, 2020).

|  | Dep: log(dwelling time) |  |  |  |
| --- | --- | --- | --- | --- |
|  | Basic model | With Epidemic-Day FE | With Stay-at-Home-Day FE | With Border Spillovers |
| log(county case) | 0.0429***<br>(0.00501) | 0.0379***<br>(0.00581) | 0.0454***<br>(0.00527) | 0.0406***<br>(0.00477) |
| log(national case) *<br>first county report | 0.00410***<br>(0.000723) | 0.00275***<br>(0.000654) | 0.00349***<br>(0.000796) | 0.00344***<br>(0.000754) |
| log(national case) | -0.00685<br>(0.00389) | -0.0179**<br>(0.00527) | -0.0100*<br>(0.00423) | -0.0253***<br>(0.00721) |
| emergency | 0.0188<br>(0.0159) | 0.0535***<br>(0.0144) | 0.0315<br>(0.0170) | 0.00720<br>(0.0134) |
| stay-home | 0.183***<br>(0.0235) | 0.151***<br>(0.0270) | 0.109***<br>(0.0279) | 0.0884<br>(0.0747) |
| school closure | 0.143***<br>(0.0100) | 0.149***<br>(0.0180) | 0.153***<br>(0.00934) | 0.133***<br>(0.0106) |
| log(county case) *<br>stay-home | -0.0237***<br>(0.00521) | -0.0213***<br>(0.00538) | -0.0311***<br>(0.00533) | -0.0258***<br>(0.00474) |
| log(total border states<br>case) |  |  |  | 0.0238**<br>(0.00683) |
| log(total border states<br>case) * stay-home |  |  |  | 0.00927<br>(0.00739) |
| R2 | 0.776 | 0.788 | 0.622 | 0.780 |
| N | 251,992 |  |  |  |

**Table 2:** Summary of regression results. The specifications are (1) models with cumulative county cases, (2) models with cumulative state rather than county cases, (3) models with cumulative state and county cases, (4) models with new county cases rather than cumulative cases, (5) models with new state cases rather than cumulative cases, (6) models with new county and state cases rather than cumulative cases, (7) models with cumulative county deaths, (8) models with cumulative state rather than county deaths, and (9) models with cumulative state and county deaths. Model (a) is the basic county fixed effects model with no time fixed effects, (b) add fixed effects relative to the first case in the county, and (c) add fixed effects relative to the date of the stay-at-home order.

| models with<br>national<br>interaction | county<br>case | state case | stay-at-home | school-closure | county<br>interaction | state<br>interaction | county crowd<br>out % | state<br>crowd out<br>% |
| --- | --- | --- | --- | --- | --- | --- | --- | --- |
| 1-a | 0.0429*** |  | 0.183*** | 0.143*** | -0.0237*** |  | 55.2% |  |
| 1-b | 0.0379*** |  | 0.151*** | 0.149*** | -0.0213*** |  | 56.2% |  |
| 1-c | 0.0454*** |  | 0.109*** | 0.153*** | -0.0311*** |  | 68.5% |  |
| 2-a |  | 0.0350** | 0.147* | 0.114*** |  | -0.00267 |  | 7.4% |
| 2-b |  | 0.0402*** | 0.204*** | 0.113*** |  | -0.0146* |  | 36.3% |
| 2-c |  | 0.0362** | 0.161* | 0.122*** |  | -0.0151 |  | 41.7% |
| 3-a | 0.0335*** | 0.0315** | 0.171** | 0.114*** | -0.0199** | -0.00304 | 59.4% | 9.5% |
| 3-b | 0.0275*** | 0.0368*** | 0.228*** | 0.114*** | -0.0135* | -0.0159* | 49.1% | 43.2% |
| 3-c | 0.0351*** | 0.0325** | 0.178** | 0.121*** | -0.0235*** | -0.0141 | 67.0% | 43.4% |
| 4-a | 0.0210*** |  | 0.166*** | 0.158*** | -0.00261 |  | 12.4% |  |
| 4-b | 0.0220*** |  | 0.124*** | 0.148*** | -0.00430 |  | 19.5% |  |
| 4-c | 0.0236*** |  | 0.0817** | 0.163*** | -0.00790 |  | 33.5% |  |
| 5-a |  | 0.0303*** | 0.219*** | 0.120*** |  | -0.0148* |  | 48.8% |
| 5-b |  | 0.0200*** | 0.171*** | 0.129*** |  | -0.0109 |  | 54.5% |
| 5-c |  | 0.0313*** | 0.154*** | 0.124*** |  | -0.0195** |  | 62.3% |
| 6-a | 0.00838 | 0.0298*** | 0.230*** | 0.120*** | 0.00718 | -0.0179* | -86.5% | 60.1% |
| 6-b | 0.0143** | 0.0189*** | 0.183*** | 0.130*** | 0.00239 | -0.0140* | -16.7% | 74.1% |
| 6-c | 0.0106 | 0.0306*** | 0.164*** | 0.124*** | 0.00323 | -0.0219** | -30.5% | 71.6% |
| 7-a | 0.0616*** |  | 0.132*** | 0.114*** | -0.0394*** |  | 64.0% |  |
| 7-b | 0.0504*** |  | 0.110*** | 0.101*** | -0.0276** |  | 54.8% |  |
| 7-c | 0.0649*** |  | 0.0751** | 0.120*** | -0.0449*** |  | 69.2% |  |
| 8-a |  | 0.0456*** | 0.168*** | 0.0809*** |  | -0.0260*** |  | 57.0% |
| 8-b |  | 0.0362*** | 0.132*** | 0.0792*** |  | -0.0172* |  | 47.5% |
| 8-c |  | 0.0471*** | 0.134*** | 0.0856*** |  | -0.0337*** |  | 71.5% |
| 9-a | 0.0227** | 0.0448*** | 0.178*** | 0.0814*** | -0.00455 | -0.0289*** | 19.8% | 64.5% |
| 9-b | 0.0238*** | 0.0352*** | 0.141*** | 0.0795*** | -0.00486 | -0.0201** | 20.2% | 57.1% |
| 9-c | 0.0240** | 0.0461*** | 0.140*** | 0.0856*** | -0.00597 | -0.0355*** | 24.6% | 77.0% |

We consider a number of additional alternatives. Replacing the natural log of time at home with the level of time at home or accounting for cases in the bordering counties or states does not qualitatively alter the results in Table 2 (SI Tables S2-S10). Including the rate of change in cases (Table S11), combining cumulative cases, new cases, and deaths (Table S12) does not qualitatively affect the estimate of the interaction between voluntary behavior and policy orders. The log of cases or the log of deaths is used because of the exponential growth of these measures. Using the arithmetic level of cases or deaths along with the time spent at home (Table S13-S15) suggests similar results with respect to cases measured at the state level, but leads to imprecise measures when cases are measured at the county level. Using time away from home as the dependent variable provides insights as the results presented in Table 2 (Tales S16-S18). Splitting counties by metropolitan and non-metropolitan areas provide a similar response to case reports and policy effects (Table S19), suggesting that certain factors like population share and transportation are well controlled in the main model specification.

The signs of all coefficients are robust to the model specification. However, the magnitude of the policy order coefficients, especially the stay-at-home order, varies across specifications suggesting that the relative influence of cases and policy on the change in time at home may be misattributed. Of specific concern is if the misattribution is shifted to (from) the direct impact of cases from (to) the stay-at-home order. There are two possible mechanisms by which an omitted time-varying variable may lead to the misattribution of causality between direct responses to case reports, which we assume are voluntary, and policy orders, which are involuntary. First, counties in which the population is more likely to voluntarily respond to the pathogen may be more likely to implement stay-at-home orders earlier. This would lead our model to overestimate the effect of the policy order. Second, county or state officials observe the voluntary response of their populations and delay orders selectively in places with a strong voluntary response. We are unaware of any public record that documents this later mechanism, and for most states, policy orders were state-wide despite heterogeneity within the state. To address this general concern, we investigate a split sample and bound the effect of potential misattribution using partial identification (29).

First, we split the sample and execute the primary specification on the 498 counties that never receive a stay-at-home order, and on the 2,606 counties that receive the stay-at-home order, with and without the interaction (Table S20). We find that a one percent increase in county cases is associated with a 1.9 percent increase in time at home, and school closures is associated with a 17 percent increase in time at home for counties without a stay-at-home order. We repeat the analysis on only the counties that experience the stay-at-home order and find that a one percent increase in county cases is associated with a 2.3 percent increase in time at home, and school closures is associated with a 14 percent increase in time at home, and the stay-at-home order increase time at home by 21 percent. This suggests that if anything, the misattribution is shifting the voluntary response effect to the policy effect, implying that our estimate of voluntary avoidance behavior is conservative.

Despite the fact that the primary pathway for misattribution suggests an overestimate of the effect of the policy order and underestimating the voluntary response, we wish to provide a conservative lower bound for the voluntary response, which limits the scope for policy to crowd out the voluntary response. First, we drop national cases and estimate the model with daily fixed effects and with state-by-day fixed effects to non-parametrically remove any temporal trends correlated nationally or within the state. This gives lower bound estimates for the response to county cases because the fixed effects likely soak up some of the voluntary response as well. We find that a one percent increase in cases is associated with a 1.9 percent and 1.8 percent increase in time at home with daily fixed effects and state-by-day fixed effects, respectively (Table S21). These values are substantially lower than those estimated in the primary specification. Next, we use a partial identification strategy and impose these levels of voluntary response so that we are replacing the assumption of no omitted confounding variable with an assumed level of voluntary response, and we re-estimate the model. With daily (state-by-day) fixed effects in the first estimate, we find that school closures increase time at home by 1.5 (1.5) percent, stay-at-home orders increase time at home by 1.6 (1.6) percent, and the interaction between stay-at-home and log of reported county cases is -0.5 (−0.5) percent. The relative magnitude of the crowding-out effect is still nearly 50 percent of the voluntary response; however, with state-by-day fixed effects, the crowding-out effect is not precisely estimated.

We are concerned about the misattribution of the mechanism of the rise in time spent at home. The assumption in any regression analysis aiming to uncover mechanisms is that there are no omitted variables correlated with an included regressor and the dependent variable. We can replace this assumption by assuming the effect of a driver that is potentially correlated with the omitted variable. This creates substantial scope for misattribution but allows us to investigate how large the misattribution would need to be to reverse the core finding -- that stay-at-home orders crowd out some of the voluntary response. We impose a range of main effects on the log of reported county cases and on the stay-at-home order to estimate the model. The misattribution would need to be substantial to discredit the finding that policy displaced and crowded out the voluntary response (Figure 3). Furthermore, Figure 3 shows that the crowding-out effect is more likely to be spurious when the voluntary response to cases or the effect of the stay-at-home order is small. This implies that if there is scope for crowding out, then there will be crowding out, and crowding out is least likely if there is not a strong effect of policy.

**Figure 3.**
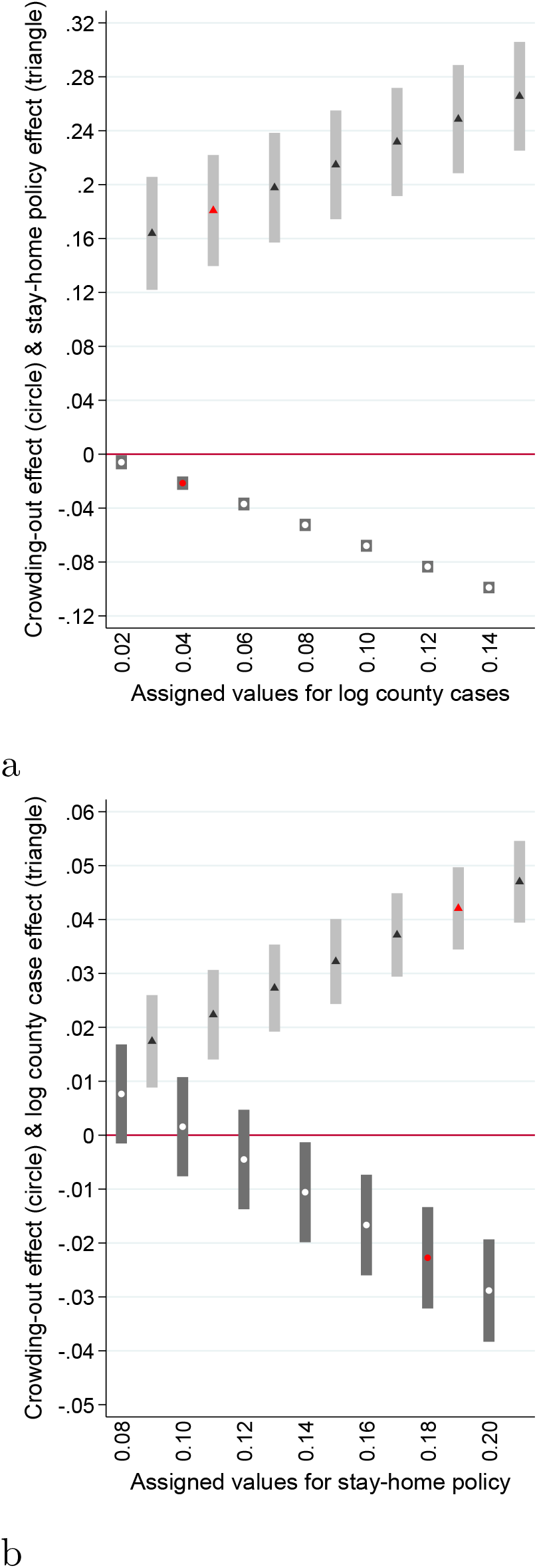
Analysis of potential misattribution using partial identification by imposing the (a) log term of county cases and (b) stay-home policy to the main effect of the other variable (triangle dots, 95CI in light gray) and estimated crowding out effect (circle dots, 95CI in dark grey). Red dots represent the real effects with the coefficients reported in Table 1 (column 1).

We focus on time at home because of its role in reducing spread. Alternatively, we can conduct a similar analysis using visits to specific types of sites (Table S22). In doing so, we find similar evidence for a crowding effect of stay-at-home orders (Figure 4).

**Figure 4.**
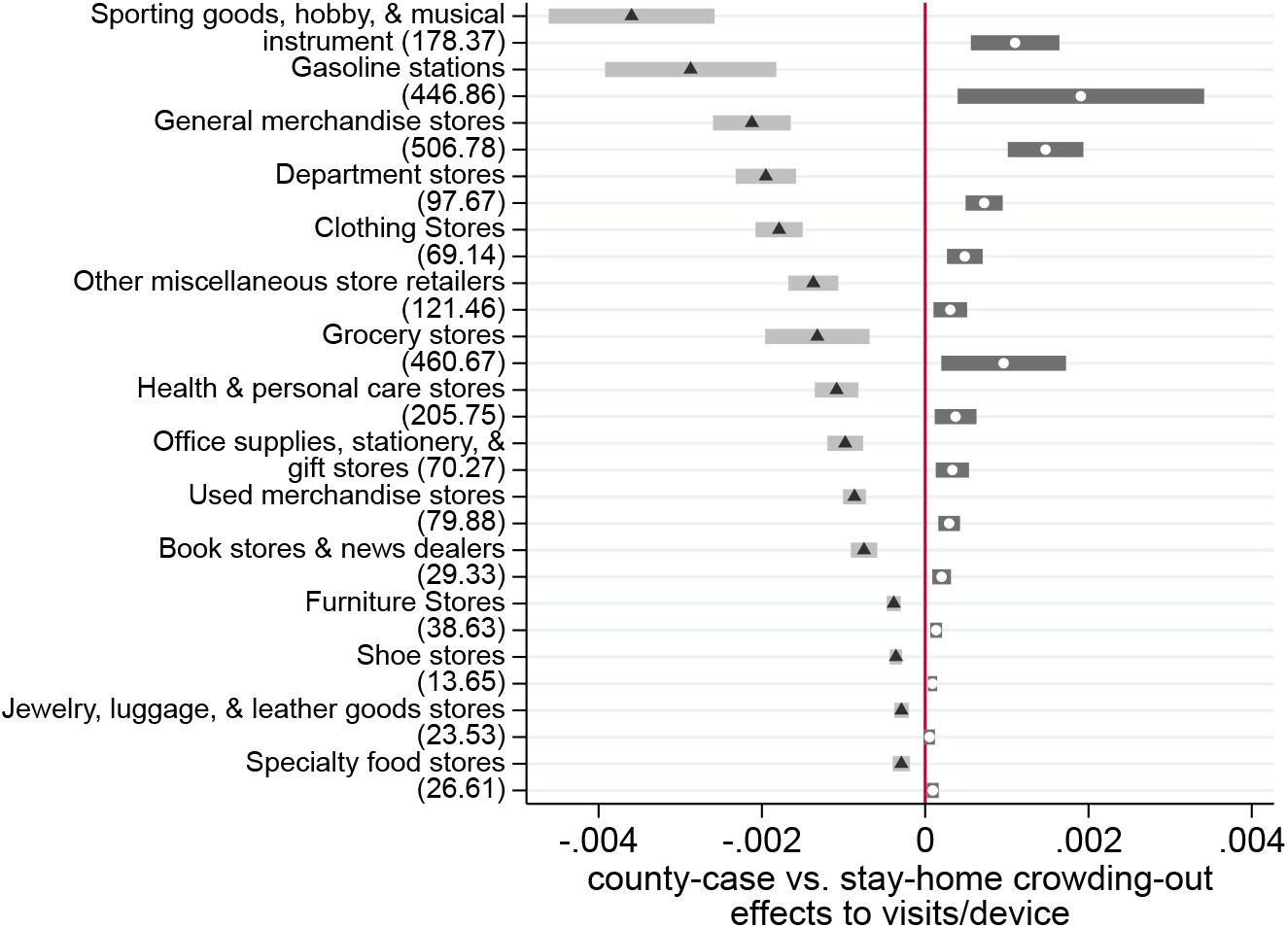
The effect of the log of county COVID-19 case reports (triangle dots, 95CI in light grey) and crowding out effect (circle dots, 95CI in dark grey) on visits to the most commonly visited retail establishments (Jan 31 – April 21, 2020). The numbers in the parentheses on the y-axis are the means of total visits for each industry group.

Americans would have altered behavior and increased time at home, at least for some period of time, in response to the increasing county, state, and national reports of COVID-19 cases, even if no orders were issued. The regression results can be used to decompose the drivers and simulate a counterfactual time at home in the absence of orders, in the absence of voluntary effects, and in the case where voluntary effects and policy orders are additively separable (Figure 5). The voluntary response alone is relatively muted, but coupled with school closures and the emergency order, it makes up a substantial share of the behavioral response.

**Figure 5.**
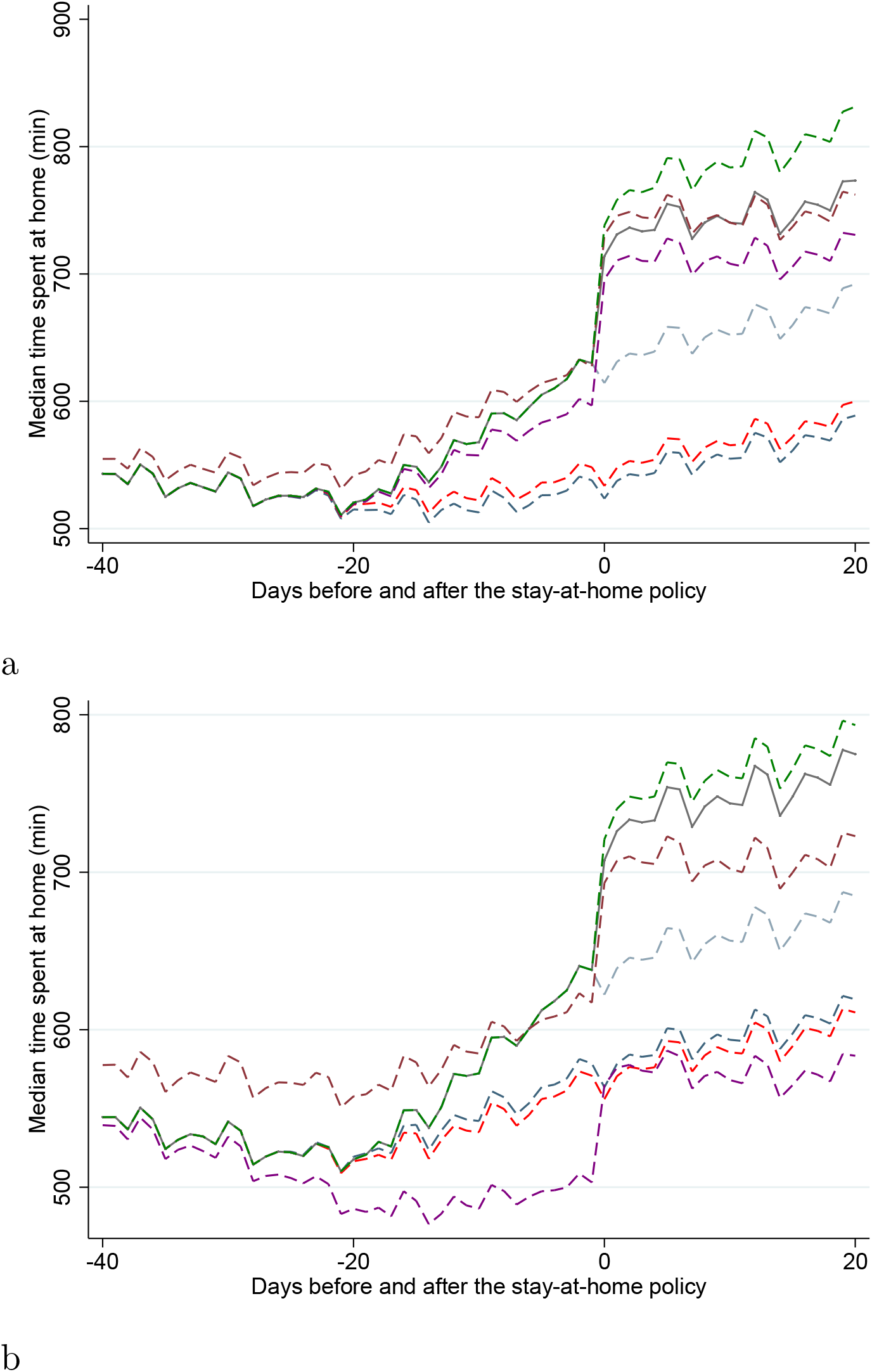
Illustration of the relative magnitude of the various contributions to staying home behavior (40 days before and 20 days after the stay-home policy), panel (a) cumulative county cases and panel is cumulative state cases. Line colors are model predicted means for (1) grey: the model predicted value; (2) blue: no state of emergency, stay-home, or school closure policies; (3) red: no stay-home or school closure policies; (4) light blue: no stay-home policy; (5) brown: no case reports; (6) purple: no county/state-case reports; (7) green: no interaction effect.

If Americans had spent less time at home, then there may have been more cases and more death, which could have led to a stronger avoidance response than the current prediction. Yet, given testing constraints, it is unlikely more cases would have been reported, and there is mounting evidence that the number of cases is many times the reported amount (30, 31).

## Discussion

Our results add to the growing evidence that Americans increased time at home during the COVID-19 epidemic and that a non-trivial share of this response was voluntary. Our results also support the growing evidence that people stayed home as the result of policies that directly and indirectly asked them to do so. There is little uncertainty among public health experts that staying home and greatly reducing contacts will slow the spread of the SARS-CoV-2 virus. What is uncertain is how much credit policy orders can take for the behavioral shifts and associated reduction in contacts in resulting pathogen spread.

Recent assessments have put all the weight on policy orders by assumption (17-19), while other analysts have focused on showing that there was a voluntary response (16). There was both, and this is unsurprising. It would be hard to believe that policy orders to stay home did not increase time at home.

What is important to understand is the relative role of policy and voluntary behavior in changing people’s behavior. Given the totality of the events, there is not enough random variation in the system to estimate and separate the three effects with perfect certainty. Nevertheless, we provide compelling evidence that a substantial share of the non-trivial voluntary response was partially replaced by policy orders.

Governments imposed restrictive policies to encourage distancing during this pandemic. After relaxing policies, many regions again face a resurgent epidemic raising the question of renewed policies like stay-at-home orders. It is important to understand the individual actions that people take to protect themselves from risk, and how they interact with public policy, in order to understand the public health benefit and tradeoffs of such policies. However, this is complicated by the fact that past experience matters and people may respond to a case level on the decline or a second wave differently than the first rise in cases.

With increased media coverage of COVID-19 morbidity and mortality along with emergency and stay at home orders, individuals may more easily recalled the adverse effects of spending too much time in public leading to voluntary social distancing. This behavior can be grounded in the availability heuristic (32). Population responses to large-scale threats are often driven by risk perception; response-efficacy, the belief that an appropriate response exists for the situation; and self-efficacy, the belief that the person can take the required action (33). Messaging that increases salience of a threat that does not emphasize a self-efficacy and response efficacy can lead to maladaptive behavior. To counter maladaptive behavior, messaging is most effective when paired with messaging that increases response-efficacy, and self-efficacy, e.g., emphasizing mortality risk is paired with actions groups and families can take (34).

Moreover, public policies may have encouraged employers to allow greater flexibility to work from home and diminish cultural pressure to simply be present in the workplace. An important side effect of school closures is that they may keep a substantial share of adults home. We did not test the salience hypothesis explicitly, and it would be difficult to separate a salience effect from a voluntary effect.

Public policies led to greater levels of time spent at home, but the effect was not completely additive to voluntary efforts. The additional time at home may be necessary since specific conditions are necessary for private behavior to fully internalize the costs of infecting others (35) or congesting hospitals. Quaas et al. (36) find that voluntary behavior was likely sufficient to contain COVID-19 but not suppress it. Still, the public measures appear to be crowding out a substantial amount of the voluntary effort. Perhaps lower cost measures exist that can lead to the same amount of time at home (or ultimately reduction in transmission) at lower cost. By highlighting the crowding-out effect, we hope to spark a search for policies that empower individuals and crowd-in voluntary contributions to controlling COVID-19. Nevertheless, a first step in accentuating voluntary behavior, crowding in rather than out, is providing information that people can respond to. In the case of COVID-19 that means local testing that provides clear, accurate, timely, and locally relevant information for people to make decisions about behavior and maintaining trust (37).

## Materials and Methods

Data on emergency declarations and stay-at-home orders are cross-referenced from five sources Julia Raifman and colleagues at Boston University (38), National Association of Counties, https://ce.naco.org/?dset=COVID-19&ind=Emergency%20Declaration%20Types the New York Times, https://www.nytimes.com/interactive/2020/us/coronavirus-stay-at-home-order.html, the COVID-19 policy trackers website https://lukaslehner.github.io/covid19policytrackers/, and the Crowdsourced COVID-19 Intervention Data, https://docs.google.com/spreadsheets/d/133Lry-k80-BfdPXhlS0VHsLEUQh5_UutqAt7czZd7ek/edit#gid=0. Reported case and death data came from the New York Times (39). All policies were put in place between Feburary 14, 2020 and April 7, 2020. District level school closure data comes from MCH Strategic Data, https://www.mchdata.com/covid19/schoolclosings. Counties were classified as metropolitan or non-metropolitan following the US Department of Agriculture Economic Research Service classification (https://www.ers.usda.gov/topics/rural-economy-population/rural-classifications/).

Data on time spent at home is based on anonymized and aggregated mobile device location data from SafeGraph (40). Benzell et al. (41) recently used these data to develop a merit order for business closing, and the use of these data is increasingly common (e.g., 42). We used three distinct SafeGraph products to quantify behavior during the epidemic. First, we used median home dwell time reported at the Census block group on each day. Dwell time is the time that a device is present at its common evening location, which is assumed to be home. Common evening location is where the device rests overnight most often over the preceding six weeks. We construct a county average of median home dwell time (Census block group) by weighting the estimate in each Census block group by the number of devices reported on that day. The normalization is required to compare estimates over time because the panel of devices changes over time, and individual Census block groups can have reporting artifacts. We dropped the lowest 1% of county dwell times to reduce the misreporting of zero or low home dwell times. Squire (43) discusses the potential biases in the SafeGraph data and provides evidence that these biases are unlikely to be large for behavior aggregated at the county level.

We construct a set of county-level weather control variables by aggregating 4km gridded estimates of maximum and minimum daily temperature, maximum and minimum relative humidity, precipitation amount, surface solar radiation (a measure of sunlight), and wind speed. Observations are by county day. The data are processed using https://github.com/jbayham/gridMETr, and based on http://www.climatologylab.org/gridmet.html (44).

We focus on the period between January 31 and April 21, 2020. As of January 31, only three travel-related cases of COVID-19 were reported in the United States. Two weeks later, on February 14, the first state of emergency declaration was issued in California. By April 21, all stay-at-home orders that were issued had been in effect for at least two weeks.

Our primary specification is

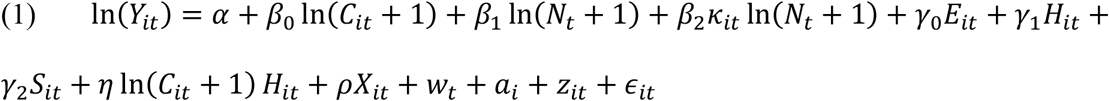

Where In (*Y*_*it*_) is the natural log of time spent at home. The case variables are *C*_*it*_, which is the cases in own county and *N*_t_, which is reported cases from the US. *κ*_*it*_ is an indicator for whether the first case in the county had occurred, which likely influences the saliency of national cases. We calculate the natural log of cases because of the exponential growth in the early phase of the epidemic. We add one to address the zeros in the data. The policy main effects are *E*_*it*_, a dummy variable indicating whether an emergency order has been issued for the county, *H*_*it*_ is a dummy variable indicating whether a stay-at-home (or equivalent shelter-in-place order) has been issued for a county, and *S*_*it*_ is a dummy variable for school closures in the county. We consider a county under a policy the first day that a county has a local or state policy. The expression In(*C*_*it*_ + 1) *H*_*it*_ is the interaction between natural log of county cases plus one and the stay-at-home order, which is the order we focus on with respect to the potential to crowd out voluntary behavior. Most counties were under emergency orders before they experienced their first case (Figure S1).

To remove potential correlation between any non-time varying omitted variable and the regressors of interest we include county fixed effects, *a*_*i*_. These fixed effects control for omitted variables such as county population and political preferences, which do not vary meaningfully over the study period. We also condition on county-level weather, *X*_*it*_ and day of week *w*_*t*_ with fixed effects. Device counts vary by county and date. Therefore, we separate device counts into 50 even size bins and include a device count fixed effect, *z*_*it*_, that varies by day and county for all models. Changing the number of device count bins between 25 and 100 does not meaningfully affect the results. We cluster standard errors at the state level to account for state-level serial correlation and heteroskedasticity caused by the phase-in orders. We choose to cluster at the state level rather than the county level because most policies are statewide.

After estimating the primary specification, we consider a number of alternative specifications. The purpose of these alternative specifications is to show that the sign of the interaction term and relative magnitudes of the main effects are stable and robust across specifications. The alternative specifications include replacing In(*Y*_*it*_) with *Y*_*it*_, including day relative to first case in the county fixed effects, day relative to stay-at-home order fixed effects, and including cases from boarding states. We replace county cases with state cases and estimate multiple specifications with both. We replace cumulative reported cases with newly reported cases at the county, state, and national level, and we estimate multiple models that have cumulative and new cases at all three levels. We replace county cases with reported deaths and we estimate models with reported cases and deaths. We replace the time at home metric with a separate metric from SafeGraph, which is time not at home. Because of the way smart device data are recorded these two metrics do not sum to 1440 minutes. Finally, we replace time at home with counts of trips to specific types of businesses that are uniquely identified in the SafeGraph Point of Interest data set. Business types are aggregated by NAICS code within a county.

Despite the stability of results across the many specification, there are two remaining potential concerns with our empirical approach. First, time spent out of the home may lead to cases leading to reverse causality. This has been a concern in research in response to other infectious agents where data were only available monthly or weekly (9). However, the time between infection, testing, and reporting is approximately 8-14 days (https://www.cdc.gov/coronavirus/2019-ncov/hcp/planning-scenarios.html#table-2). This lag breaks the potential for simultaneity within the daily data.

The second concern with our empirical approach is that county cases are correlated with when governments enact orders in a time-varying manner – if this correlation is not time-varying, then county fixed effects remove the confound. However, if counties that are more or less likely to spend time at home in response to cases are more or less likely to adopt policies earlier or later, then the estimated impact of the policy could be biased. If the story were that counties where the population is more likely to respond voluntarily are also more likely to enact a policy early, then we expect the bias to lead to an overestimate of the effect of policy and an underestimate of the voluntary response. The converse story is that policymakers observe their citizens voluntarily distancing and delay orders, which would reverse the misattribution. Either way, it is unclear what this misattribution would mean for the interaction effect needed to identify policy orders crowding out voluntary behavior.

Addressing this concern to the level of point identification requires finding a way to remove this unobserved potential trend in a system with a large number of treated units (45). Given, the magnitude of the events around COVID-19, the credibility of any instrument would be challenged. We employ three approaches to bound the potential for the omitted confound to lead us qualitatively astray, building confidence in our core results. The first approach splits the sample into counties that never experience a stay-at-home order and the subset of counties that do. While the counties that never experience a stay-at-home order are not ideal controls for the other counties, the more credible omitted variables story suggests that these counties should show a more muted voluntary responsive than counties that experienced orders, and we expect that under the confounding trend hypothesis they provide a lower bound on voluntary behavior.

Next, we employ two partial identification strategies. By employing a partial identification strategy we concede that point identification may not be feasible, but investigate how large misattribution would need to be to undermine our core findings (29).

First, we consider the possibility that the temporal confound operates at the national or state level. We estimate the response to reported county cases with day and with state-by-day fixed effects. This non-parametrically removes common national or state-level time trends. National or state cases are perfectly correlated with the day fixed effects, so we do not include them at this stage. We argue that this is a lower bound on the response to county cases because the daily fixed effects likely capture some of the direct response to cases as more cases are reported. We then impose the lower bound estimate for the behavioral response to county case reports and re-estimate the main specification, conditional on the imposed, and assumed response to county case reports. This would not address a dynamic county-level confound. Therefore, we replace the assumption that no such confound exists, which is implicit in our primary specification, with an assumed range of main effects for effect for county cases or the stay-at-home order and re-estimate the model. This allows us to investigate the range of main effects over which the interaction term remains negative, lending credibility to the crowding out findings.

## Data Availability

Data used for this analysis are publicly available from Safe Graph, New York Times, and COVID-19 US state policy database maintained by colleagues at Boston University.

## Acknowledgments

Youpei Yan and Eli P. Fenichel are supported by the Knobloch Family Foundation and the Tobin Center for Economic Policy Analysis and Amyn A. Malik and Saad B. Omer are supported by the Yale Institute for Global Health. We acknowledge support from Amazon Web Service Diagnostic Development Initiative.

